# How do household living conditions and gender-related decision-making influence child stunting in Rwanda? A population-based study

**DOI:** 10.1101/2023.08.19.23294315

**Authors:** Jean N. Utumatwishima, Ingrid Mogren, Aline Umubyeyi, Ali Mansourian, Gunilla Krantz

**Affiliations:** University of Gothenburg; University of Rwanda; Umea University; Lund University

## Abstract

Child stunting (chronic undernutrition) is a major public health concern in low- and middle-income countries. In Rwanda, an estimated 33% of children are affected. This study investigated the household living conditions and the impact of gender-related decision-making on child stunting. The findings contribute to ongoing discussion on this critical public health issue. In December 2021, a population-based cross-sectional study was conducted in Rwanda’s Northern Province; 601 women with children aged 1–36 months were included. Stunting was assessed using low height-for-age criteria. The Multidimensional Poverty Index (MPI) was used to determine household socioeconomic status. Researcher-designed questionnaires evaluated gender-related factors such as social support and household decision-making. Multivariable logistic regression analysis identified risk factor patterns. Six hundred and one children were included in the study; 27.1% (*n*=163) were diagnosed as stunted; there was a higher prevalence of stunting in boys (60.1%) than girls (39.9%; p<0.001). The MPI was 0.265 with no significant difference between households with stunted children (MPI, 0.263; 95% confidence interval [CI], 0.216–0.310) and non-stunted children (MPI, 0.265; 95% CI, 0.237–0.293). Most households reported a lack of adequate housing (78.9%), electricity (63.0%), good water sources (58.7%), and proper toilets (57.1%). Male-headed households were predominant (92% vs 8.0%; *p*=0.018), although women often shared decision-making with their partners; 26.4% of the women reported they were forced to have sexual intercourse within marriage (*p*=0.028). Lack of support during illness (odds ratio [OR], 1.93; 95% CI, 1.13–3.28) and absence of personal guidance (OR, 2.44; 95% CI, 1.41–4.26) were significantly associated with child stunting (*p*=0.011). Poverty contributes to child stunting in the Northern Province of Rwanda. Limited social support and women’s lack of decision-making power in the household increase stunting rates. Interventions should empower women and address the broader social and economic context to promote both women’s and children’s health.

## Introduction

Undernutrition has been studied extensively in different contexts in low- and middle-income countries (LMICs) [1–5]. The most consistent factors associated with childhood undernutrition in sub-Saharan Africa are as follows: women’s low education level, mothers’ age <20 years, women’s low body mass index (BMI) (<18.5 kg/m^2^), fathers’ low level of education, rural residence, unimproved source of drinking water, poor household wealth; further factors related to the child were low birth weight, increasing age, male sex, and episodes of diarrhea [6–8]. Different interventions to address undernutrition have been initiated and in the last decades, the global undernourishment rates have declined, which has improved the health and nutritional status in children <5 years [9]. However, there is an imbalance between the desired level of improvement in undernutrition and the nutritional interventions actually implemented [10]. Studies undertaken in sub-Saharan Africa demonstrate that the decrease in undernutrition is slow, and improvements are distributed unequally in different areas within the same country [11]. For example, families living in the northern region of Ghana had higher rates of underweight children compared with other regions due to poor socioeconomic conditions [12]. In a review that included data from Demographic and Health Surveys (DHS) in 35 LMIC countries, socioeconomic conditions were the strongest factors associated with child anthropometric failures [13]. This explains why some countries initiated several interventions to reduce poverty [14]. Stunting is an outcome of chronic undernutrition that reflects poverty and failure to receive adequate nutrition over a longer period [10–12].

The global Multidimensional Poverty Index (MPI), created using the multidimensional measurement method of Alkire and Foster, estimates the poverty level of a country and provides details of poverty deprivations across different regions of the same country [15]. In Rwanda, based on the 2019/2020 DHS data, the MPI calculated at national level was 0.231 [16]. The MPI ranges between 0 and 1, and a higher MPI indicates a greater level of poverty within a region [16].

Rwanda, like other LMICs, has implemented various programs to combat malnutrition, including providing each family with one cow, offering micro-nutrients to vulnerable children, as well as providing school feeding programs across the country [15]. Despite equal distribution of resources and initiatives, stunting remains a significant problem mainly in the Western and Northern Provinces of Rwanda, where 40% and 41%, respectively, of children <5 years are stunted compared with the city of Kigali, and the Eastern and Southern Provinces, where the rates are 21%, 29%, and 33%, respectively [11]. According to the Rwanda DHS reports of 2014/2015 and 2019/2020, stunting rates in other provinces have decreased, whereas the prevalence of stunting in the Northern Province has increased slightly [14, 17].

In addition, recent evidence shows that women’s health and their position in the household are critical in relation to children’s nutritional status [18, 19]. Gender refers to the roles, attributes, behaviors, and opportunities associated with being male, female, or gender non-binary [18]. Men and women display distinct health behaviors, encounter diverse health vulnerabilities, and receive varying responses from health systems. Currently, the prevalence of stunting in Rwanda is 33%. Despite the Rwandan government’s extensive efforts to address stunting and promote gender equality in the country, no study has been conducted to examine the gender-related factors linked to stunting in Rwanda.

This research program forms part of the collaboration between the University of Rwanda and five Swedish universities, titled “Undernutrition, an interdisciplinary program focused on children and their mothers”. It is designed as an interdisciplinary epidemiological study with qualitative components, divided into four different research projects. The aim of the program is to investigate the chronic problem of stunting by combining theories and methods from different fields of science (agriculture/veterinary science, public health, obstetrics, pediatrics, nutrition, geographical information systems, innovation), and to establish a pilot version of a Rwandan Medical Birth Register. The aim of this study was to investigate household living conditions and the impact of gender-related decision-making on child stunting.

## Methods

### Study design, study population, and sample size

A cross-sectional study was conducted on a sample of children aged from 1 month to 3 years and their mothers who were representative of the population. The sample size was calculated based on the prevalence of stunting in the population, with a desired precision of 4% and a 95% confidence interval (CI). The final sample consisted of 630 households randomly selected from all five districts of the Northern Province. During data collection, mothers in 29 households failed to provide the necessary data for some of the analyses, giving a participation rate of 95.4% (*N*=601).

Multi-stage random sampling was used to identify households for inclusion. The selection of the primary sampling units (villages), households, and study participants in the five districts was made in three steps. First, 186 villages were randomly selected from the total number of 2743 existing villages using the geospatial grid system. Second, households in each village were selected proportionate to the total number of households in each village. Third, a mother with a child aged 1–36 months of age was asked for an interview.

The community health worker responsible for the village listed all households with eligible mothers, and this list constituted the village sampling frame. The first participant to be interviewed was identified using a generated random number. Then, a calculated sampling interval in each village was applied to get the next household, depending on the number of households to be selected in each village. If the eligible person in the household was absent, the closest household on the list was selected, assuming that the living conditions between neighboring households would be similar. The selection of households also took into consideration the need for a random selection of villages and the spread of measurement points for the Geographical Information System and spatial analyses to be performed by collaborators within the same project.

### Data collection procedures

A comprehensive questionnaire was developed to capture a range of factors associated with undernutrition, such as sociodemographic and psychosocial factors, maternal and child health characteristics, access to food, anthropometric measures, crops, animal husbandry, and milk quality. The questionnaire was translated into Kinyarwanda, the national language in Rwanda. The questions used in this study were validated in previous studies [20–22]. The questionnaire was then implemented as an Android-based app on the emGeo platform to facilitate digital data collection and storage on a centralized cloud-based database.

The University of Rwanda, College of Medicine and Health Sciences, School of Public Health was the lead implementer of the survey. A group of 13 experienced interviewers comprising six PhD students, eight registered nurses, and one post-doc conducted the survey under the supervision of Rwandan and Swedish supervisors. Five days of pre-field training took place followed by 2 days to pilot the questionnaire in one selected village in the Northern Province. The data collection took place in December 2021.

### Measurement of dependent variables

Stunting was defined as low height-for-age, which is measured as less than −2 standard deviations of the median value of the international growth reference. The distribution of height and weight among children between 1 and 36 months was compared against the WHO Child Growth Standards reference population [23]. Weight, height, and age data were used to calculate height-for-age z scores based on the World Health Organization’s anthropometric references [24]. Standard techniques were used to measure the child’s weight and height. Length was measured in a recumbent position in children <2 years and in a standing position for children > 2 years using a ShorrBoard measuring board. Weight measurements were taken using SECA scales with a digital display (model number SECA 874). Participating children were then categorized as stunted or not stunted.

### Measurement of independent variables

#### Factors related to the mother and her partner

Sociodemographic and psychosocial factors were categorized and analyzed as independent risk factors. The age of the mother was captured as a continuous variable and later categorized into three categories (≤20, 21–34, and ≥35 years of age). BMI, defined as the mother’s weight in kilograms divided by the square height in meters [21], was categorized into three groups: underweight (≤18.49 kg/m^2^), healthy weight (18.50–24.9 kg/m^2^), and overweight (25.0–29.9 kg/m^2^) (6). Marital status included five categories: cohabiting with a man/woman, divorced or separated, married in monogamy or polygamy, single mother, and widow. Marital status was dichotomized into married (or cohabitating) and living alone (being single, divorced, widowed, or separated). Educational level for both the mother and her partner consisted of any school attendance. It was categorized into incomplete primary school, complete primary school, secondary school or more. Occupation was categorized into skilled work (civil servants and students) and non-skilled work (farmers, vocational training).

#### Factors related to the children

Age was measured as a continuous variable in months and categorized into three groups: 1–12 months, 13–24 months, and 25–36 months. Birth weight was categorized into five groups: ≤2499 g, 2500–2999 g, 3000–3499 g, 3500–3999 g, and ≥4000 g. Sex of the child was dichotomized into girls and boys. Places of birth were at home, primary health facility (health center or health post), and secondary health facility (district hospitals and other referral hospitals).

#### Factors related to the household

Sex of the head of household was grouped into male or female. Mothers’ marital status was categorized into married and other relationships (daughter, sister, or any other close person). Household income per month was categorized into three categories (≤17,500, 17,501–35,999, and ≥36,000 Rwandan francs).

### Women’s living conditions and household assets

Core welfare indicators are key social indicators for population sub-groups. Developed jointly by the World Bank with UNDP and UNICEF, the Core Welfare Indicators Questionnaire is designed to monitor social indicators in sub-Saharan Africa on an annual basis [25]. Core welfare indicators such as the type of the house, the quality of the water source, access to electricity, distance traveled to reach an improved water source, cooking facilities, quality of toilet facilities, and assets in the household were dichotomized as improved or not improved. Assets in the household included the availability of a mobile phone, radio, television, refrigerator, bicycle, motorcycle, and a car. The composite assets index was categorized into two groups: those with no assets or only one asset, and those with two or more assets. To complete the assessment of living conditions, the global MPI was calculated for the 601 households. The MPI is an international measure of acute multidimensional poverty. It complements traditional poverty measures by capturing the acute deprivations in health, education, and living standards that a person faces simultaneously [26].

### Social support and women’s power in the family

The Social Support Questionnaire-Short Form, a 6-item measure of social support, was used [27]. For every question, the options were “sometimes”, “often”, “always”, and “never”. The responses were dichotomized into yes (for “sometimes”, “often”, and “always” answers, equal to the reference category) and no (for “never” answers, equal to the exposure category). Social support was defined as having a friend or family member who would assist in case of illness or would share food, housing, lend money, or offer psychological guidance during problem times.

Women’s power at household level was assessed using the United Nations Economic Commission questionnaire for the decision-making process within households alongside selected items from the Women’s Empowerment Questionnaire of the United Kingdom Data Archive [28, 29]. The following household decisions were assessed: major purchases, family visits, use of earned money, seeking personal healthcare, access to family planning, consensual sexual intercourse, and condom use during sexual intercourse. The options for these types of decisions were “joint decision”, “personal decision”, “partner’s decision”, and “other people’s decisions”. In addition, home and land ownership were assessed and recorded as joint ownership, personal ownership, or no ownership.

### Statistical analysis

Prevalence (number and %) was used to describe participants’ characteristics. The dependent variable was child stunting, dichotomized into stunted and not stunted. The distribution of sociodemographic and psychosocial factors and test of difference for two groups were calculated by applying Pearson’s chi-squared test. Associations between child stunting and perceived social support were estimated by multivariable logistic regression models. *p* values ≤0.05 were considered statistically significant. All measures of association are presented as odds ratios (ORs) with their 95% CI.

The calculation of the MPI involves assigning equal weights to each dimension and equally weighting each indicator within a dimension. The following stepwise methodology was used to calculate the MPI [30]. A person’s deprivation status was determined based on a cutoff, and their overall deprivation score was calculated by adding up the weights of the indicators in which they were deprived. The global MPI in this study considered seven indicators, excluding child mortality, school attendance, and cooking fuel because data on these indicators were not available in the dataset. The MPI calculation involved aggregating the dimensions and indicators using the adjusted headcount ratio formula.

All analyses were performed using STATA 17 SE (College Station, TX, USA).

### Ethical considerations

The research protocol and study tools were approved for scientific and ethical integrity by the Institutional Review Board (IRB) of the University of Rwanda, College of Medicine and Health Sciences (review approval notice no. 181/CMHS IRB/2021) and the National Health Research Committee (no. NHRC/2020/PROT/047). The participant consent form described the purpose of the study, that participation in the study was voluntary and that refusal or withdrawal at any stage in the research process would involve no penalty or loss, currently or in the future. The interviewers obtained written consent from the respondents before the interview. The consent form explained how anthropometrics would be estimated. It was further explained that in the event of withdrawal, all data related to the woman and her child would be deleted. Participants were informed of their freedom to choose whether to answer questions, including sensitive topics such as partner violence, mental health, and child abuse. The benefits of the study for the mother, child, household, community, and country were discussed. If any participant expressed a need for health care intervention, they were referred to a nearby health center or hospital. In cases where severe illness was identified during the interview, both the mother and child were assisted in seeking health care at the nearest health facility.

## Results

### Sociodemographic characteristics

This study included women aged between 18 and 58 years, distributed across age groups; 51.7% of the children were girls and 48.3% were boys. Of the 601 children, 27.1% (*n*=163) were diagnosed with stunting, and the prevalence of stunting was higher among boys than girls (60.1% vs 39.9%, *p*<0.001) (Table 1). Marital status, women’s level of education, and occupation were equally distributed between the stunted and non-stunted groups, as were partner’s age, level of education, and occupation (Table 1). Most household heads were male (92% vs 8%, *p*=0.018), and most households (57.4%) earned less than 17,500 RWF (∼17 USD) per month, with no clear difference observed between the stunted and non-stunted groups. House and land ownership were jointly shared between mothers and their partners in most families and were not associated with child stunting. However, stunted children were more likely to be found in households without a lactating cow (79.1% vs 20.9%).

**Table 1.**
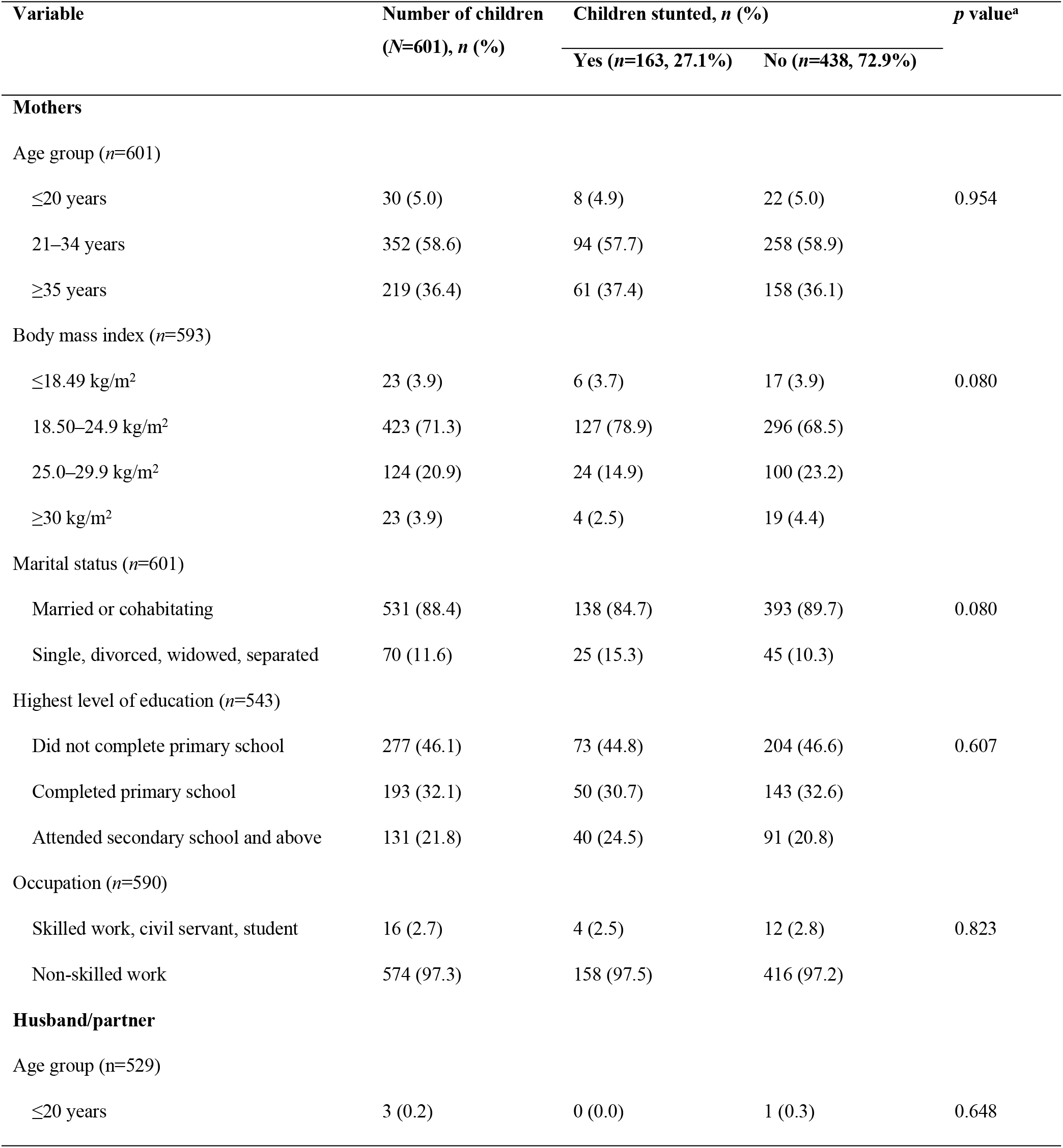
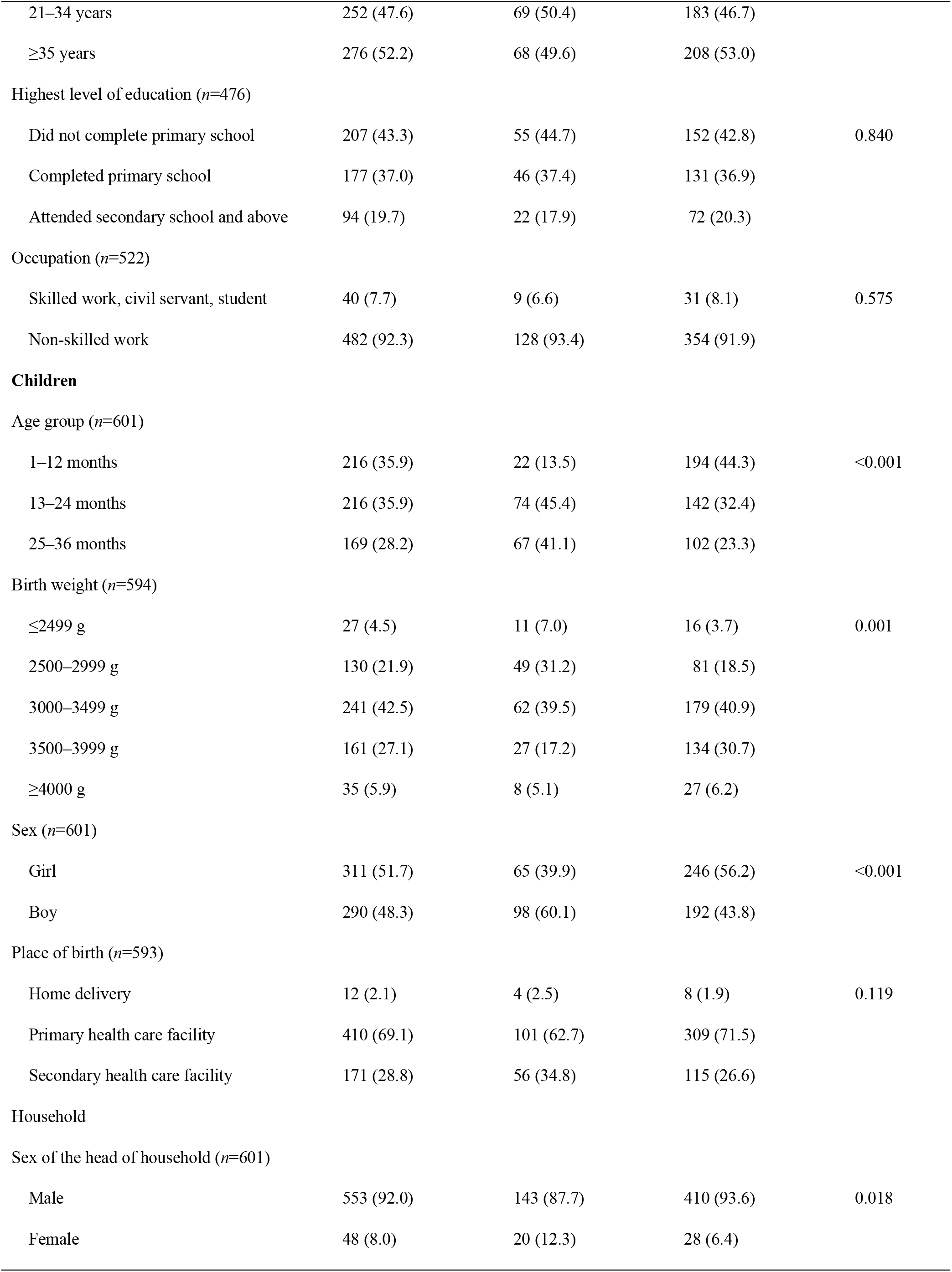
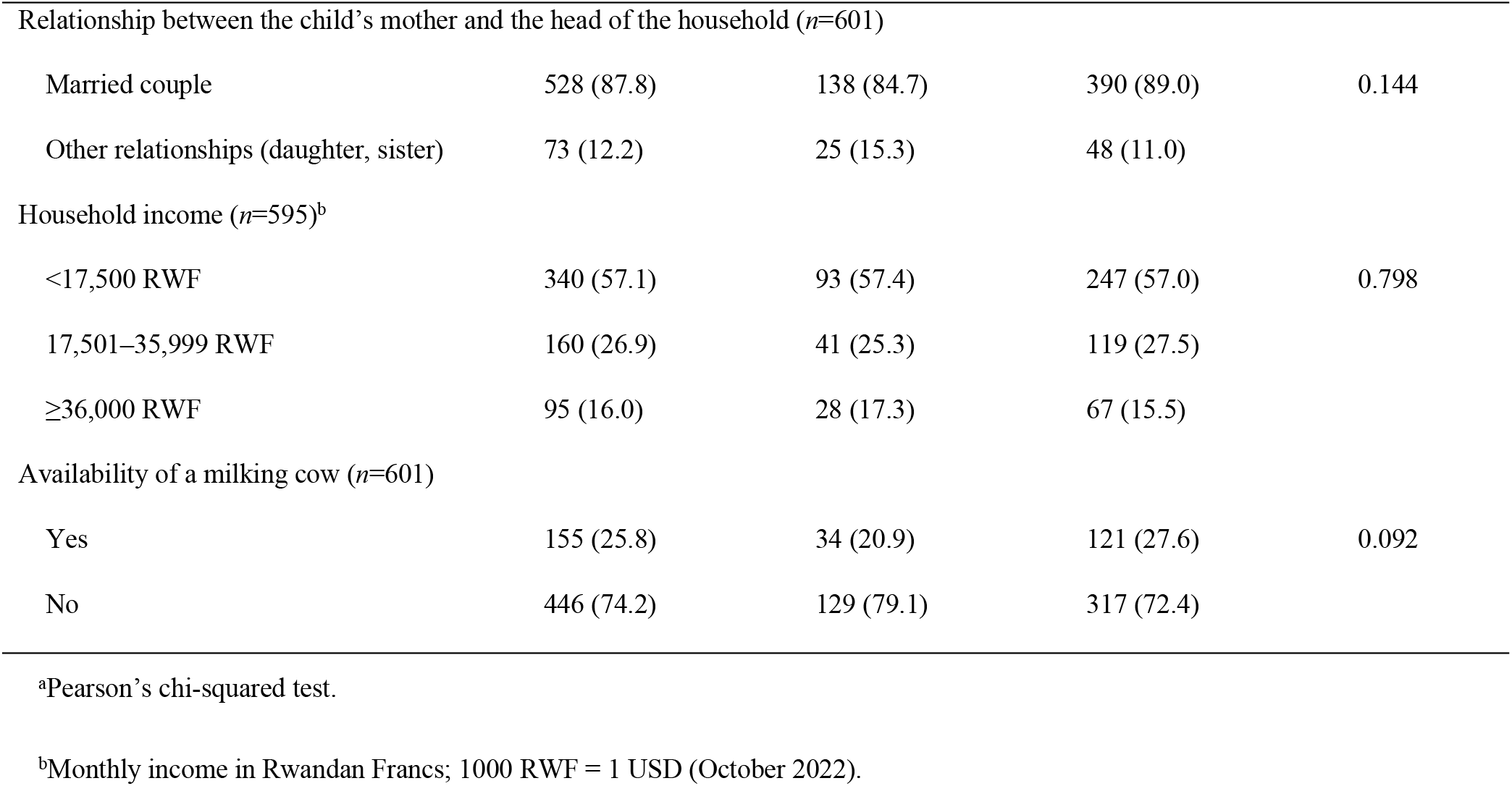
Sociodemographic and psychosocial background factors of the study population.

The mean weight of mothers of stunted children was significantly lower (55.9±8.6 kg) than that of mothers of non-stunted children (58.9±8.5 kg) (*p*<0.001). Similarly, the study showed that the mean height of mothers of stunted children was 155.7±8.5 cm, and that of mothers of non-stunted children was 157.7±8.5 cm, which was statistically significant (*p*=0.010) between the two groups. Parity was correlated with stunting (*p*=0.027); the highest prevalence of stunting was observed among children born to mothers with five or more children. Furthermore, a greater proportion of mothers with stunted children (40.4%) reported poor self-rated health compared with mothers of non-stunted children (31.0%). Stunted children had statistically significant lower birth weight (3049±526 g) than non-stunted children (3246±484 g) (*p*<0.001). In addition, stunted children were older (22.4±8.4 months) compared with non-stunted children (16.1±9.9 months) (*p*<0.001). Higher rates of diarrhea and fever in the 2 weeks before the survey were found in stunted children compared with non-stunted children (29.2% vs 22.1% and 33.1% vs 29.1%, respectively). The prevalence of stunting was significantly higher among non-breastfed children (34.4%) compared with breastfed children (20.1%) (*p*<0.001). Moreover, the prevalence of mothers’ reporting poor health status in their children (19%) was higher compared with the non-stunted group (11.4%) (*p*=0.014).

### Living conditions, household assets, and Multidimensional Poverty Index

With regard to households, 78.4% lived in unimproved houses, and only 21.6% had improved houses (Table 2). Of the children living in households in unimproved houses, 80.0% were stunted, whereas only 20.0% were stunted among those living in households in improved houses. About 56.7% of households did not have improved toilet facilities (traditional toilet facility without modern hygiene and sanitation features) and in these households, the prevalence of stunting was 62.3%; in households with improved toilets (sewage, flushing, and other amenities), the prevalence of stunting was 37.7%, the difference was not statistically significant (*p*=0.097). Lack of access to electricity in the household (62.2%) was associated with 2.09 times higher odds of children being stunted compared with households with access to electricity, even after adjusting for household monthly income, education of the mother and the partner (adjusted OR, 2.09; 95% CI, 1.34–3.28). The prevalence of stunting was higher among households with one or no assets (67.9 %) compared with those with two or more assets (32.1%), and the difference was close to statistical significance (*p*=0.051) (Table 2).

**Table 2.** Living conditions of the women and household assets (*N*=601)

| Variable | Number of children<br>(*N*=601), *n* (%) | Children stunted, *n* (%) |  | *p* value<sup>a</sup> |
| --- | --- | --- | --- | --- |
|  |  | Yes (*n*=163, 27.1%) | No (*n*=438, 72.9%) |  |
| Type of house (*n*=573) |  |  |  |  |
| Improved | 124 (21.6) | 31 (20.0) | 93 (22.3) | 0.561 |
| Unimproved | 449 (78.4) | 124 (80.0) | 325 (77.7) |  |
| Water source (*n*=578) |  |  |  |  |
| Improved | 246 (42.6) | 63 (39.9) | 183 (43.6) | 0.423 |
| Unimproved | 332 (57.4) | 95 (60.1) | 237 (56.4) |  |
| Access to electricity (*n*=598) |  |  |  |  |
| Yes | 226 (37.8) | 41 (25.3) | 185 (42.4) | <0.001 |
| No | 372 (62.2) | 121 (74.7) | 251 (57.6) |  |
| Distance to water source >30 min walk (*n*=581) |  |  |  |  |
| No | 346 (59.5) | 93 (58.9) | 253 (59.8) | 0.836 |
| Yes | 235 (40.5) | 65 (41.1) | 170 (40.2) |  |
| Toilet facilities (*n*=591) |  |  |  |  |
| Improved | 256 (43.3) | 60 (37.7) | 196 (45.4) | 0.097 |
| Not improved | 335 (56.7) | 99 (62.3) | 236 (54.6) |  |
| Assets available in the household |  |  |  |  |
| Radio (*n*=598) | 243 (40.6) | 57 (35.2) | 186 (42.7) | 0.098 |
| Mobile phone (*n*=598) | 513 (85.8) | 134 (82.7) | 379 (86.9) | 0.190 |
| Television set (*n*=598) | 22 (3.7) | 2 (1.2) | 20 (4.6) | 0.053 |
| Refrigerator (*n*=598) | 1 (0.2) | 0 (0.0) | 1 (0.2) | 0.542 |
| Bicycle (*n*=593) | 70 (11.8) | 16 (10.0) | 54 (12.5) | 0.408 |
| Motorcycle (*n*=593) | 8 (1.4) | 0 (0.0) | 8 (1.9) | 0.080 |
| Car (*n*=593) | 1 (0.2) | 1 (0.6) | 0 (0.0) | 0.100 |
| Composite Assets index, (*n*=598) |  |  |  |  |
| ≥2 assets and more | 230 (38.5) | 52 (32.1) | 178 (40.8) | 0.051 |
| No asset or one asset only | 368 (61.5) | 110 (67.9) | 258 (59.2) |  |

<sup>a</sup>Pearson's chi-squared test.

The MPI among households with stunted children was 0.263 (95% CI, 0.216–0.310; standard error, 0.024) (Table 3). Conversely, households without stunted children had an MPI of 0.265 (95% CI, 0.237–0.293; standard error, 0.014). The highest deprivation rates were found in the living conditions domain; 78.9% lacked improved houses and 63.0% lacked access to electricity. In addition, 58.7% lacked improved water sources and 57.1% lacked improved toilets. The deprivation rate for the education domain was 44.9% for the education of mothers or their partners, and assets had a deprivation rate of 37.2%.

**Table 3.**
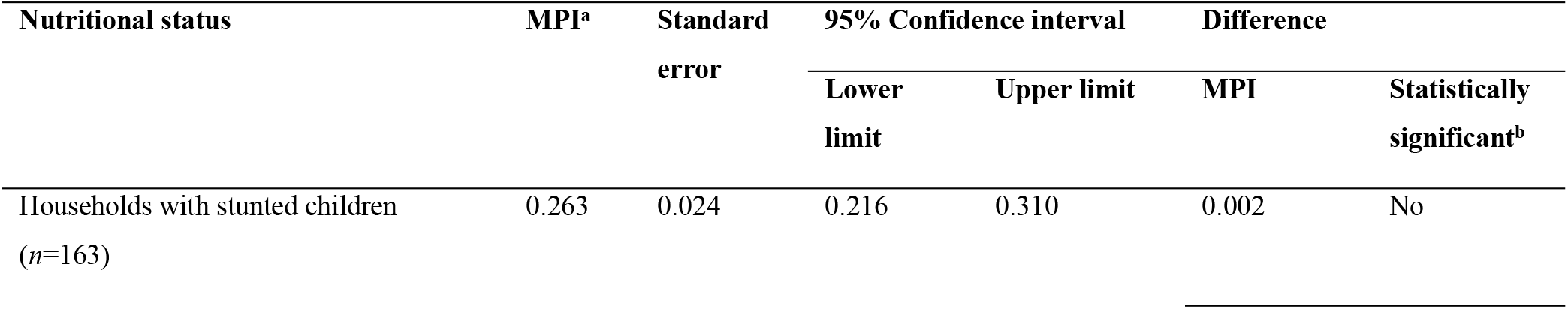
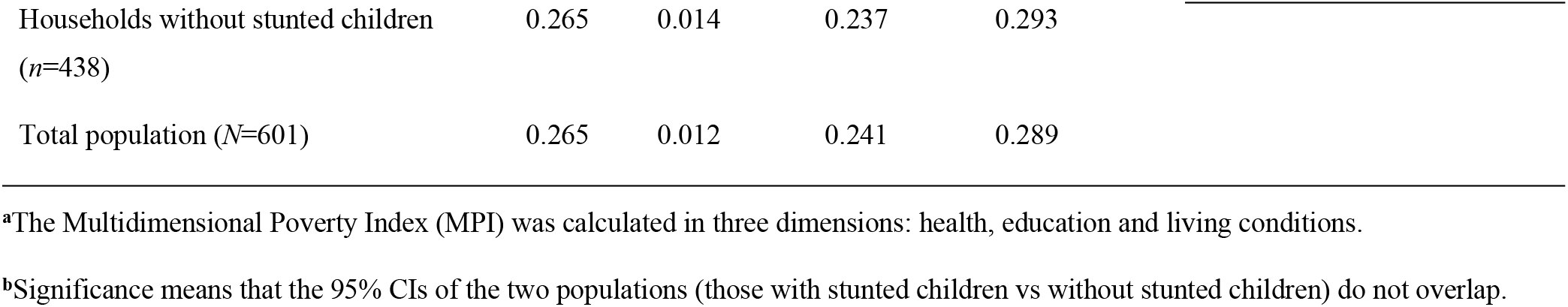
Calculation of the Multidimensional Poverty Index at household level (*N*=601)

| Nutritional status | MPI<sup>a</sup> | Standard error | 95% Confidence interval |  | Difference |  |
| --- | --- | --- | --- | --- | --- | --- |
|  |  |  | Lower limit | Upper limit | MPI | Statistically significant<sup>b</sup> |
| Households with stunted children (*n*=163) | 0.263 | 0.024 | 0.216 | 0.310 | 0.002 | No |

|  |  |  |  |  |
| --- | --- | --- | --- | --- |
| Households without stunted children<br>(*n*=438) | 0.265 | 0.014 | 0.237 | 0.293 |
| Total population (*N*=601) | 0.265 | 0.012 | 0.241 | 0.289 |

<sup>a</sup>The Multidimensional Poverty Index (MPI) was calculated in three dimensions: health, education and living conditions.
<sup>b</sup>Significance means that the 95% CIs of the two populations (those with stunted children vs without stunted children) do not overlap.

### Social support and child stunting

Households in which children were diagnosed with stunting exhibited a lower distribution of social support for all six types. In the initial crude logistic regression analyses (Table 4), “lack of support during illness” (OR, 2.01; 95% CI, 1.29–3.13) and “not having someone to provide guidance during personal problems” (OR, 1.88; 95% CI, 1.19–3.00) were statistically significant risk factors for children’s exposure to stunting. After adjusting for covariates, including age, education, income, and household leadership, lack of support during illness (adjusted OR, 1.93; 95% CI, 1.13–3.28) and lack of guidance during personal problems (adjusted OR, 2.44; 95% CI, 1.41–4.26) remained statistically significant and associated with child stunting.

**Table 4.** Women perceived social support and association with child stunting (*N*=601)

| Type of support | Number of children<br>(*N*=601), *n* (%) | Children stunted, *n* (%) |  | Odds ratio (95% confidence interval) |  |
| --- | --- | --- | --- | --- | --- |
|  |  | Yes (*n*=163, 27.1%) | No (*n*=438, 72.9%) | Crude<sup>a</sup> | Adjusted<sup>b</sup> |
| Support during illness (*n*=594) |  |  |  |  |  |
| Yes | 488 (82.1) | 120 (74.1) | 368 (85.2) | 1 |  |
| No | 106 (17.8) | 42 (25.9) | 64 (14.8) | **2.01 (1.29–3.13)** | **1.93 (1.13–3.28)<sup>3</sup>** |
| Sharing food (*n*=600) |  |  |  |  |  |
| Yes | 390 (65.0) | 98 (60.1) | 292 (66.8) | 1 |  |
| No | 210 (35.0) | 65 (39.9) | 145 (33.2) | 1.34 (0.92–1.94) | 1.15 (0.74–1.77) |
| Sharing a house (*n*=599) |  |  |  |  |  |
| Yes | 350 (58.4) | 85 (52.5) | 265 (60.6) | 1 |  |
| No | 249 (41.6) | 77 (47.5) | 172 (39.4) | 1.39 (0.97–2.01) | 1.40 (0.92–2.13) |
| Borrowing money (*n*=600) |  |  |  |  |  |
| Yes | 328 (54.7) | 79 (48.5) | 249 (57.0) | 1 |  |
| No | 272 (45.3) | 84 (51.5) | 188 (43.0) | 1.41 (0.98–2.02) | 1.29 (0.85–1.96) |
| Guidance during problems (*n*= 600) |  |  |  |  |  |
| Yes | 507 (84.5) | 127 (77.9) | 380 (87.0) | 1 |  |
| No | 93 (15.5) | 36 (22.1) | 57 (13.0) | **1.88 (1.19–3.00)** | **2.44 (1.41–4.26)<sup>3</sup>** |
| Tangible support for personal problems (*n*=599) |  |  |  |  |  |
| Yes | 465 (77.6) | 118 (72.4) | 347 (79.6) | 1 |  |
| No | 134 (22.4) | 45 (27.6) | 89 (20.4) | 1.49 (0.98–2.25) | 1.44 (0.89–2.34) |

Values in bold type are statistically significant.
<sup>a</sup>Multivariable logistic regression with 95% confidence interval.
<sup>b</sup>Values are controlled for age of mothers and children, education level of mother and partner, sex of the head of the household, and household income.

### Household decision-making and child stunting

Joint decisions between mothers and their partner/husband were most prevalent across all decision-making domains. Nonetheless, a statistically significant difference was observed between households with stunted and non-stunted children regarding the mother’s decision-making autonomy in major household purchases (*p*=0.004), visits to family and friends (*p*=0.016), and utilization of earned money (Table 5). Specifically, 36% of the households were single-mother households, and a higher rate of stunting was found among the children in these households compared with households with two parents. Conversely, there were no statistically significant differences in joint decisions and decisions made by the husband/others between households with stunted and non-stunted children. Households with mothers who experienced forced sexual intercourse (*n*=152, 26.4%) (Table 5) had a statistically significantly higher prevalence of stunting (33.1%) compared with those who did not report such events (24.0%) (*p*=0.028).

**Table 5.** Status of the women’s household decision-making capacity (*N*=601)

| Participation in household decision-making | Number of | Children stunted | *p* value<sup>a</sup> |
| --- | --- | --- | --- |

|  | children (N=601) | Yes (n=163,<br>27.1%) | No (n=438,<br>72.9%) |  |
| --- | --- | --- | --- | --- |
| Major household purchases (n=585) |  |  |  |  |
| Mother's decision only | 51 (8.7) | 23 (14.6) | 28 (6.6) | 0.004 |
| Joint decision with partner/husband | 281 (48.0) | 78 (49.4) | 203 (47.5) |  |
| Husband's decision/others | 253 (43.3) | 57 (36.1) | 196 (45.9) |  |
| Visits to family and friends (n=584) |  |  |  |  |
| Mother's decision only | 83 (14.2) | 33 (20.9) | 50 (11.7) | 0.016 |
| Joint decision with partner/husband | 353 (60.5) | 91 (57.6) | 262 (61.5) |  |
| Husband's decision/others | 148 (25.3) | 34 (21.5) | 114 (26.8) |  |
| Utilization of earned money (n=582) |  |  |  |  |
| Mother's decision only | 53 (9.1) | 23 (14.7) | 30 (7.1) | 0.015 |
| Joint decision with partner/husband | 323 (55.5) | 85 (54.1) | 238 (56.0) |  |
| Husband's decision/others | 206 (35.4) | 49 (31.2) | 157 (36.9) |  |
| Personal health care (n=585) |  |  |  |  |
| Mother's decision only | 263 (45.0) | 71 (45.2) | 192 (44.9) | 0.869 |
| Joint decision with partner/husband | 228 (39.0) | 59 (37.6) | 169 (39.5) |  |
| Husband's decision/others | 94 (16.0) | 27 (17.2) | 67 (15.6) |  |
| Family planning (n=583) |  |  |  |  |
| Mother's decision only | 40 (6.9) | 11 (7.0) | 29 (6.8) | 0.349 |
| Joint decision with partner/husband | 157 (26.9) | 49 (31.2) | 108 (25.4) |  |
| Husband's decision/others | 386 (66.2) | 97 (61.8) | 289 (67.8) |  |
| Experienced forced sexual intercourse (n=575) |  |  |  |  |
| Yes | 152 (26.4) | 51 (33.1) | 101 (24.0) | 0.028 |
| No | 423 (73.6) | 103 (66.9) | 320 (76.0) |  |
| Condom use (n=573) |  |  |  |  |
| Yes | 301 (52.5) | 71 (46.4) | 201 (47.9) | 0.758 |
| No | 272 (47.5) | 82 (53.6) | 219 (52.1) |  |
| Land ownership (n=586) |  |  |  |  |
| Personal land | 20 (3.4) | 45 (28.3) | 107 (25.1) | 0.268 |
| Joint ownership with husband | 414 (70.7) | 106 (66.7) | 308 (72.1) |  |
| No land owned | 152 (25.9) | 8 (5.0) | 12 (2.8) |  |
| House ownership (*n*=585) |  |  |  |  |
| Personal house | 11 (1.9) | 6 (3.8) | 5 (1.2) | 0.114 |
| Joint ownership with husband | 452 (77.3) | 119 (75.3) | 333 (78.0) |  |
| No house owned | 122 (20.8) | 33 (20.9) | 89 (20.8) |  |

<sup>a</sup>Pearson's chi-squared test.

## Discussion

This study, conducted in the Northern Province of Rwanda, found that child stunting is a huge problem affecting children >1 year, and boys more often than girls. Furthermore, the MPI revealed that poverty is the primary cause of child stunting. Poor living conditions, such as the lack of electricity, were identified as a contributing factor to child stunting. Lack of decision-making power for women regarding consensual sexual intercourse, as well as inadequate support systems, are also factors that increase the likelihood of stunting in children.

### Prevalence of child stunting

According to the 2019/2020 Rwanda Demographic and Health Survey (RDHS) [14], the prevalence of stunting among children <5 years old in the Northern Province was 41%, which is higher than the prevalence found in this study (27.1% among children <3 years). However, the RDHS included children up to 5 years old, whereas this study focused only on children ages 1–36 months. In addition, the prevalence of stunting in Rwanda has been declining in recent years, as indicated by data from the RDHS. A study conducted in rural Ethiopia reports a stunting prevalence of 47.9% among children aged 6–59 months, and a study conducted in Nigeria reports a prevalence of 37% among children <5 years of age [23, 24]. These differences in prevalence could be attributed to variations in socioeconomic status, dietary habits, access to health care, and other factors specific to each population, but still indicate the scale of the problem in other sub-Saharan African countries.

### Child health and stunting

Rwanda has made significant efforts to reduce mortality in children <5 years through various interventions, including increasing the number of antenatal care visits, involving community health workers, and implementing community-based health insurance schemes [31]. However, low birth weight and hygiene-related diseases remain prevalent in the country [32]. In our study, we found that birth weight was lower for stunted children compared with non-stunted children. Moreover, stunted children had a higher incidence of diarrhea and fever in the past 2 weeks and a greater likelihood of not being breastfed. These findings are consistent with previous studies showing that low birth weight (<2500 g) is a risk factor for stunting and other health issues later in life [33–37]. The connection between stunting, diarrhea, and fever is also well established in the literature, likely attributed to inadequate nutrition and poor living conditions that can compromise a child’s immune system [38]. Furthermore, our findings reinforce the notion that breastfeeding is associated with a lower incidence of stunting because breast milk offers optimal nutrition for infants and is critical for growth and development [39]. Children who are not breastfed or who are breastfed for a short time are at an increased risk of stunting and other health problems [40].

### Mother’s health and child stunting

We also found that the weight and height of mothers of stunted children were significantly lower compared with mothers of non-stunted children. This finding is consistent with other studies that have shown a relationship between maternal nutrition and stunting in children [41–43]. Therefore, improving maternal nutrition is a critical component in addressing stunting in children. In addition, parity was associated with stunting; the prevalence of stunting was highest among children of mothers with five or more children. A study conducted in Indonesia reported that children born to mothers with higher parity (defined as five children or more) were more likely to be stunted than children born to mothers with lower parity [44]. Studies in Ethiopia and Indonesia found a similar association between parity and increased risk of child stunting [44, 45]. These findings suggest that there may be a cumulative effect of the number of children on maternal and child health outcomes. It highlights the importance of providing adequate antenatal care and nutrition education to women, particularly those with a higher number of children.

Mothers of stunted children were more likely to rate their overall health as poor compared with mothers of non-stunted children. This finding is particularly concerning because maternal health plays a critical role in child health outcomes [7, 46]. Maternal health interventions should therefore be prioritized to improve the health of mothers and their offspring. From a longer-term perspective, poverty should be alleviated, women should be better educated on how to feed their children, and as a result, many children will be healthy and consequently contribute to a reduction in overall family size.

### Living conditions and child stunting

Poor living conditions, characterized by lack of assets and electricity in the household, expose children to stunting. The lack of household assets places mothers under constant stress, potentially reducing their feeding capacity and resulting in less time for child care [33]. Furthermore, children living in households without improved sanitation were more likely to be stunted than children living in households with improved sanitation. This finding has been corroborated in other studies conducted in LMICs that established the association between unimproved sanitation and stunting [35–37]. The findings of this study indicate a higher presence of radios and electricity in households with non-stunted children. The radio plays a vital role in communicating best practices for improving sanitation, as demonstrated in a study conducted in Tanzania [38]. Families lacking electricity tend to lack a radio as well, leading to a lack of crucial information on children’s health priorities, exacerbating the malnutrition problem in the community.

The MPI score based on population survey data in the Northern Province (0.263) was higher than the national MPI (0.231) calculated using the 2019/2020 DHS data by the Oxford team in 2022 [16]. Similarity can be observed by comparing the stunting rate with the MPI index. Data from the 2019/2020 DHS indicate the following stunting rates and their corresponding MPIs: Northern Province 41% (MPI, 0.264), Western Region 40% (MPI, 0.264), Southern Region 33% (MPI, 0.251), Eastern Region 29% (MPI, 0.232), and Kigali city 21% (MPI, 0.100) [16, 47]. The findings from the Oxford group regarding MPI calculations in Rwanda and stunting estimates from the Rwanda DHS confirm that the MPI mirrors the stunting situation in a region. The higher the MPI, the higher the prevalence of stunting in the region. Regarding the deprivation profile, the lack of improved housing and electricity are the most dominant issues at the national level, as observed in the Northern Province as well.

### Social support

Households with children diagnosed with stunting showed a lower incidence of social support for all six types of support. Lack of support during illness and not having someone to provide guidance during personal problems were identified as statistically significant risk factors for children’s exposure to stunting. These findings are consistent with previous research that has demonstrated the importance of social support in promoting child health and development. For instance, a study in Uganda found that social support from family and community members was positively associated with child nutritional status [48]. Similarly, a study in Bangladesh found that maternal support was positively associated with child growth [49]. The present study adds to this body of research by providing evidence of the specific types of social support that are most strongly associated with child stunting. Specifically, lack of support during illness and lack of guidance during personal problems emerged as the most significant risk factors. These findings suggest that interventions aimed at improving child health and nutrition in Rwanda should focus on increasing social support for families, particularly during times of illness and personal stress.

### Household decision-making and childhood stunting

Autonomy of women can be defined in various ways; it generally refers to a woman’s ability to make decisions that influence herself and her family in her context [33]. Such decisions may include family matters, finances, work, social events, health care, travel, family planning, and childcare. In this study, most household decisions were reported to be made jointly by men and women in cohabiting couples, whereas in households headed by women, the decisions were solely made by the woman. Several studies conducted in sub-Saharan African countries link women’s decision-making power with the nutritional status of children [34]. In the current study, women’s personal decisions related to household expenses, family visits, and the use of their earnings did not protect children from malnutrition. This finding supports a study conducted in the Democratic Republic of the Congo, where female power at home did not have a positive impact on child stunting [35]. Women who lived alone, including single mothers and divorced women living in poor socioeconomic conditions, were constantly stressed and unable to provide adequate food for their children, even if they were able to make decisions. However, women who were denied the right to choose whether to have sexual intercourse tended to have stunted children. This could be because they risked becoming pregnant with unwanted children that they could not raise happily.

### Methodological considerations

This study utilized a representative sample of mothers with children between 1 and 36 months of age selected from all districts in the Northern Province of Rwanda, using of spatial grid techniques. Validated questions were used to investigate social support and women’s household decision-making capacity. Data collection (i.e. interviews) were conducted by experienced PhD students and licensed nurses with extensive interviewing experience. Although the study’s strength lies in its comprehensive sampling approach, it is a cross-sectional study, which by definition limits the interpretation of findings regarding the direction between different gender dimensions and living conditions at the household level and the nutritional status of children. Further studies are needed to support the causal relationship between household decision-making and social support on the nutritional status of children in the Rwandan context. The sensitive nature of some of the information gathered may have led to under-reporting of household difficulties faced by women, indicating that the results are not overestimated. Furthermore, the participation rate of 95.4% achieved during data collection indicates a noteworthy strength of this study. However, it should be acknowledged that in 29 households, mothers were unable to provide essential data required for specific analyses, which may have somewhat affected the representativeness of the sample.

## Conclusion

Childhood stunting remains a challenging public health problem in LMICs and an indicator of human capital development that is rooted in poverty. This study in Rwanda revealed a higher prevalence of stunting in households with low social support for women. Even when women reported decision-making power within the household, it did not have a positive impact on their children’s nutritional status. Therefore, multifaceted efforts to eliminate childhood stunting must address social and economic support for mothers in poor households as well as nutritional interventions that target the child as the primary beneficiary.

## Data Availability

All relevant data are within the manuscript and its Supporting Information files.

## Acknowledgments

We would like to express our gratitude to all the mothers and children who participated in the study and graciously shared their life experiences with us. We also extend our thanks to the interviewers, community health workers, and all others involved in the study for their invaluable assistance in completing this study.

## Notes

### Competing Interest Statement

The authors have declared no competing interest.

### Funding Statement

JN received a sponsorship from the Swedish International Development Cooperation Agency (SIDA). https://www.sida.se/en. The funders had no role in study design, data collection and analysis, decision to publish, or preparation of the manuscript.

### Author Declarations

The research protocol and study tools were approved for scientific and ethical integrity by the Institutional Review Board (IRB) of the University of Rwanda, College of Medicine and Health Sciences (review approval notice no. 181/CMHS IRB/2021) and the National Health Research Committee (no. NHRC/2020/PROT/047)

